# Genetically predicted expression of cuproptosis and copper-transport genes and coronary artery disease risk: a cis-Mendelian randomization and colocalization study

**DOI:** 10.64898/2026.09.03.26362207

**Authors:** Wang Guan, Li Wanqi, Guan Meizuo, Xing Yue

## Abstract

**Background:** Cuproptosis, a copper-dependent form of regulated cell death, has been repeatedly reported to be transcriptionally activated in atherosclerotic plaque, and cuproptosis- and copper-transport genes have been nominated as candidate biomarkers and therapeutic targets on the basis of descriptive expression studies. Whether genetically determined expression of these genes is *causally* related to atherosclerotic cardiovascular disease has not been tested.

**Methods:** We first confirmed, across two independent carotid plaque transcriptomic cohorts (GSE43292 and GSE28829), that a panel of copper-metabolism and cuproptosis genes is concordantly dysregulated in plaque and that the copper importers track the degree of inflammatory-cell infiltration. We then performed two-sample cis-Mendelian randomization (MR) and Bayesian colocalization to test the causal effect of the genetically predicted expression of 11 cuproptosis/copper genes on coronary artery disease (CAD). Instruments were cis-eQTLs from whole-blood (eQTLGen, n = 31,684) and tibial artery (GTEx v8, n = 584); the outcome was CAD (CARDIoGRAMplusC4D/Aragam 2022; 181,522 cases, 1,165,690 participants, European ancestry). Large-artery atherosclerotic stroke (GIGASTROKE) was examined as a secondary outcome. *LPL* served as a positive control. The positive control was intended to validate the direction and harmonization of the MR pipeline, not to demonstrate that single-causal-variant colocalization must succeed at a known multi-signal locus such as *LPL*.

**Results:** In plaque, copper-import genes (*SLC31A2*, *SLC31A1*) were upregulated and correlated strongly with myeloid/inflammatory infiltration (*SLC31A2* vs. M1-macrophage r = +0.87), whereas the exporter *ATP7B* correlated inversely (r = −0.73). The positive control *LPL* showed the expected robust protective effect on CAD (IVW OR 0.90 per SD higher expression, 95% CI 0.87–0.93, P = 4.1 × 10^−12^), validating the pipeline. None of the 11 cuproptosis/copper genes showed colocalization-supported causal evidence for CAD. Two genes reached nominal MR significance in single-instrument analyses (*MTF1* OR 1.16, P = 2.7 × 10^−3^; *DLAT* OR 1.41, P = 2.2 × 10^−3^) but were not supported by colocalization (*MTF1* PP.H4 = 4 × 10^−6^ with PP.H3 = 0.9998; *DLAT* PP.H4 = 0.29), indicating linkage-disequilibrium confounding rather than causality. For well-instrumented genes (e.g. *ATP7B*, *SLC31A2*, *FDX1*), estimates were null and precise enough to exclude odds ratios larger than ≈1.05 per SD.

**Conclusions:** Common cis-regulatory variation determining the baseline expression of cuproptosis/copper-transport genes in available eQTL tissues did not support a detectable causal effect on CAD risk. This does not exclude locally acquired plaque cuproptosis; rather, the reported upregulation of these genes in plaque is most consistent with a consequence or marker of established disease rather than a germline-encoded causal driver, and cautions against target nomination based on descriptive transcriptomics alone.

## 1. Introduction

Atherosclerotic cardiovascular disease remains the leading cause of death worldwide, and despite effective lipid-lowering therapy a substantial residual risk persists, motivating the search for new causal pathways and druggable targets. Copper is an essential redox-active trace element, and disturbances of copper homeostasis have long been linked— observationally—to vascular disease. Cellular copper is tightly controlled by dedicated importers (*SLC31A1*/CTR1, *SLC31A2*/CTR2), exporters (*ATP7A*, *ATP7B*), and chaperones, which together buffer the metal between essential enzymatic use and toxic accumulation.

In 2022, Tsvetkov and colleagues defined *cuproptosis*, a distinct, copper-dependent form of regulated cell death.^1^ Mechanistically, excess intracellular copper binds directly to lipoylated components of the tricarboxylic-acid cycle—chiefly the dihydrolipoamide *S*-acetyltransferase *DLAT*—causing their aggregation, proteotoxic stress and cell death; the mitochondrial ferredoxin *FDX1* acts as an upstream regulator of protein lipoylation and is required for the process, with *LIAS*, *LIPT1*, *DLD*, *DLST*, *PDHA1* and *DBT* constituting the associated lipoylation/TCA machinery.^1^ Because macrophage death and defective efferocytosis are central to necrotic-core formation in plaque, cuproptosis is an attractive candidate mechanism in atherosclerosis.^2^

Prompted by this mechanistic appeal, a large and rapidly growing bioinformatic literature has reported that cuproptosis- and copper-related genes are differentially expressed in atherosclerotic tissue, and has nominated several of them—most often *SLC31A2*, *FDX1* and *ATP7A*—as diagnostic biomarkers or candidate therapeutic targets, typically via differential-expression analysis of public microarray datasets combined with machine-learning feature selection and immune-infiltration correlation.^3,4,5^ We reproduce the core of this observation here (Section 3.1).

These studies, however, are almost exclusively *associative*: they demonstrate that gene expression differs between plaque and control tissue, but cannot establish whether that difference contributes to disease or merely reflects it. The distinction is not academic. Copper handling is pharmacologically tractable—chelators such as trientine and D-penicillamine are in routine clinical use for Wilson disease, which is caused by loss-of-function *ATP7B* mutations—so a genuine causal role for copper metabolism in atherosclerosis would justify repurposing or target-development efforts, whereas a purely reactive association would not, and might waste them. Moreover, expression changes measured in end-stage diseased tissue are especially vulnerable to reverse causation, because the inflamed, hypoxic, macrophage-rich plaque itself remodels the transcriptome.

Mendelian randomization (MR) addresses exactly this problem. It uses germline genetic variants as instrumental variables to estimate the causal effect of an exposure on an outcome, and—because genotype is fixed at conception and randomized at meiosis—is largely protected from reverse causation and environmental confounding.^6,7^ When the exposure is the cis-regulated expression of a specific gene (cis-eQTL MR), the resulting estimate approximates the effect of lifelong modulation of that gene, analogous to pharmacological target perturbation.^8,9^ Cis-MR has one important vulnerability—an instrument may tag a nearby causal variant of a *different* gene through linkage disequilibrium—which statistical colocalization directly addresses by testing whether the eQTL and disease association signals share a single causal variant.^10,11^ Used together, cis-MR and colocalization can separate genes whose expression causally influences disease from bystanders that merely lie in an associated locus.

Here we perform, to our knowledge, the first cis-eQTL Mendelian randomization and colocalization analysis of cuproptosis- and copper-transport gene expression in relation to coronary artery disease (CAD) risk. We anchor the study in our own two-cohort observation that these genes are upregulated in carotid plaque in proportion to inflammatory infiltration, and then interrogate causality with cis-MR and colocalization across two eQTL sources and a well-powered European CAD GWAS, using the lipid gene *LPL* as a positive control to validate the analytic pipeline and an explicit power analysis to quantify what effect sizes our null results can exclude.

## 2. Methods

### 2.1 Gene panel

We studied 15 genes spanning copper transport (*SLC31A1*, *SLC31A2*, *ATP7A*, *ATP7B*) and the core cuproptosis / lipoylation pathway (*FDX1*, *LIAS*, *LIPT1*, *DLD*, *DLAT*, *DLST*, *PDHA1*, *DBT*, *MTF1*, *GLS*, *CDKN2A*). *LPL* (lipoprotein lipase), a lipid gene with a well-established causal relationship to CAD, was analysed as a positive control to validate the pipeline; *APOE* was included where instruments were available.

### 2.2 Transcriptomic motivation (observational)

Two publicly available carotid atherosclerosis microarray datasets were used to establish that the panel genes are dysregulated in plaque: **GSE43292** (Ayari & Bricca 2013; 32 carotid atheroma plaque vs. 32 paired macroscopically intact tissue; Affymetrix Human Gene 1.0 ST, GPL6244)^12^ and **GSE28829** (Döring et al. 2012; 16 advanced vs. 13 early carotid plaque; Affymetrix U133 Plus 2.0, GPL570).^13^ Differential expression was assessed with a moderated *t*-test using empirical-Bayes variance shrinkage (a Python implementation equivalent to the *limma* eBayes procedure), with Benjamini–Hochberg control of the false discovery rate; genes were called differentially expressed at adjusted P < 0.05 and |log_2_ fold-change| > 0.5, applied identically to both cohorts. Probe-level results were collapsed to gene symbols. Cross-cohort concordance of the direction of effect was assessed for the copper/cuproptosis panel.

Over-representation analysis of the plaque-upregulated gene set was performed by Fisher’s exact test against the GO Biological Process (2023) and KEGG (2021, human) libraries, using all annotated platform genes as background and Benjamini–Hochberg correction. Immune-cell infiltration was estimated per sample in GSE43292 by single-sample gene-set enrichment analysis (ssGSEA; *gseapy*,^14^ rank normalization), with immune-cell marker sets from Bindea et al.^15^ and M1/M2 macrophage markers as previously defined.^16,17^ Pearson correlations between panel-gene expression and infiltration scores were computed across the 64 samples.

### 2.3 Instruments (exposures)

Cis-eQTLs were obtained from two sources:

- **Whole blood — eQTLGen** (n = 31,684),^18^ the primary instrument source because of its large sample size. For colocalization the full cis-eQTL summary statistics were used; for instrument selection the genome-wide-significant subset was used. eQTLGen reports Z-scores; we converted these to effect sizes using the accompanying allele-frequency file with the standard formulae β = Z / √[2·MAF·(1−MAF)·(N + Z^19^)] and SE = 1 / √[2·MAF·(1−MAF)·(N + Z^19^)]. The assessed allele was treated as the effect allele. **Conversion was verified by confirming that β/SE reproduced the reported Z-score for all variants (agreement 100%).**
- **Tibial artery — GTEx v8** (n = 584),^20^ a disease-relevant tissue, used in a parallel analysis.

For each gene, cis-eQTLs at P < 5 × 10^−8^ within ±1 Mb of the gene body (GENCODE gene coordinates) were clumped to independent instruments (r^2^< 0.001, 10-Mb window) using PLINK v1.90^21^ with the 1000 Genomes Phase 3 European reference panel.^22^ Instrument strength was quantified by the F-statistic; variants with F < 10 were excluded.

### 2.4 Outcomes

The primary outcome was coronary artery disease (CARDIoGRAMplusC4D/Aragam et al. 2022; GWAS Catalog accession **GCST90132314**; 181,522 cases and 984,168 controls, all of European ancestry).^23^ The secondary outcome was large-artery atherosclerotic stroke (GIGASTROKE/Mishra et al. 2022; accession **GCST90104542**; 6,399 European cases, 1,234,808 controls),^24^ selected because its pathology—large-artery plaque—most closely matches the carotid tissue of the transcriptomic analysis. All datasets used GRCh38 coordinates; genome-build concordance between exposure and outcome was verified by rsID-matched position comparison (100% agreement).

### 2.5 Statistical analysis

Exposure and outcome datasets were harmonized by rsID. Strand orientation was verified empirically rather than assumed: across all cis regions the effect allele of the outcome GWAS matched the ALT allele of the eQTL dataset in >99.8% of shared variants with zero allele flips, establishing that both datasets are reported on the GRCh38 forward strand with ALT as the effect allele. Palindromic (A/T, C/G) variants are therefore not strand-ambiguous in this setting and were retained (harmonization action 1); excluding them would have discarded ∼15% of variants per region, including the lead CAD variant of the *FDX1* locus. For genes with ≥3 instruments, the primary estimate was the inverse-variance-weighted (IVW) method, complemented by MR-Egger,^25^ weighted median,^26^ and weighted mode; heterogeneity was assessed by Cochran’s Q and directional pleiotropy by the MR-Egger intercept. Genes with a single instrument were analysed by the Wald ratio. All Mendelian randomization and colocalization analyses were performed in R v4.6.1 using *TwoSampleMR* v0.7.9,^27^ *ieugwasr* v1.1.0, *coloc* v5.2.3 (default priors p1 = p2 = 1 × 10^−4^, p12 = 1 × 10^−5^), and *data.table* v1.18.4; instrument clumping used local PLINK (via *genetics.binaRies* v0.1.2) against the 1000 Genomes European reference panel. Transcriptomic differential-expression, over-representation and single-sample gene-set enrichment analyses were performed in Python 3 using NumPy, pandas, SciPy, statsmodels, Matplotlib and *gseapy*.^31^

Bayesian colocalization (*coloc*, single-causal-variant assumption)^10^ was performed for each gene using all cis-region variants shared between the eQTL and outcome datasets (not only genome-wide-significant variants), with the default prior probabilities p1 = 1 × 10^−4^, p2 = 1 × 10^−4^ and p12 = 1 × 10^−5^. A posterior probability of a shared causal variant (PP.H4) > 0.7 was taken as evidence of colocalization; a high PP.H3 indicates distinct causal variants for the two traits, and a high PP.H1 indicates an eQTL signal with no detectable outcome association in the region.

Statistical power was summarized as the minimum odds ratio per SD of expression detectable at 80% power and α = 0.05, given each gene’s instrument strength (variance explained by the instruments and outcome case/control counts), following the standard two-sample MR power approximation. Multiple testing was addressed by a Bonferroni threshold of 0.05/14 = 3.6 × 10^−3^.

## 3. Results

### 3.1 Copper/cuproptosis genes are concordantly dysregulated in carotid plaque

Applying identical criteria to both cohorts (adjusted P < 0.05 and |log_2_ fold-change| > 0.5), differential expression analysis identified 1,307 dysregulated genes in GSE43292 (plaque vs. intact; 725 upregulated, 582 downregulated in plaque) and 909 in GSE28829 (advanced vs. early plaque; 591 up, 318 down). Focusing on the copper/cuproptosis panel, all 14 genes assessed in both datasets showed a concordant direction of dysregulation between the two independent cohorts (14/14 concordant). *SLC31A2*/CTR2 was upregulated in plaque in both cohorts (log_2_ fold-change +0.63 in GSE43292 and +0.99 in GSE28829; mean expression 8.69 in plaque vs. 8.06 in intact tissue in GSE43292), as were the copper importer *SLC31A1*, the cuproptosis regulator *FDX1*, and the exporter *ATP7B* (**Figure 1**).

**Figure 1.**
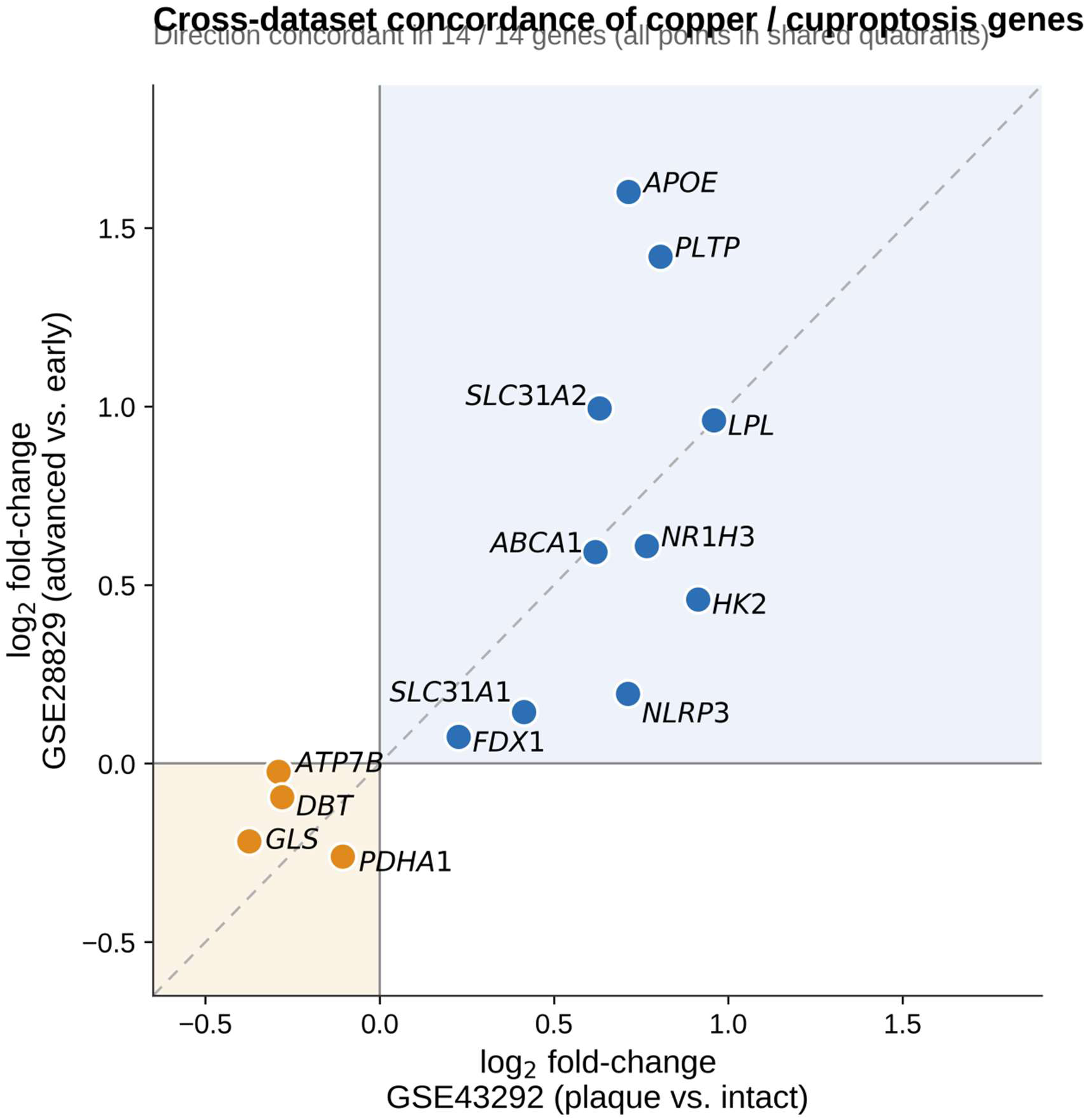
Copper- and cuproptosis-gene dysregulation in carotid plaque across two independent cohorts. Log₂ fold-change for each panel gene in GSE43292 (plaque vs. paired macroscopically intact carotid tissue, n = 32 pairs) and GSE28829 (advanced vs. early plaque, 16 vs. 13). All 14 genes assessed in both datasets show a concordant direction of change.

Functional enrichment of the plaque-upregulated genes was dominated by inflammatory and myeloid programs rather than by copper- or metabolism-specific terms. The most strongly enriched Gene Ontology biological processes were *inflammatory response* (adjusted P = 1.0 × 10^−18^), *positive regulation of MAPK cascade* (2.3 × 10^−16^), *regulation of the ERK1/ERK2 cascade* (1.4 × 10^−15^) and *cellular response to cytokine stimulus* (8.6 × 10^−12^); the top KEGG pathways were *neutrophil extracellular trap formation* (4.3 × 10^−13^), *chemokine signaling* (8.4 × 10^−13^), *cell adhesion molecules* and *phagosome* (**Figure 2**).

**Figure 2.**
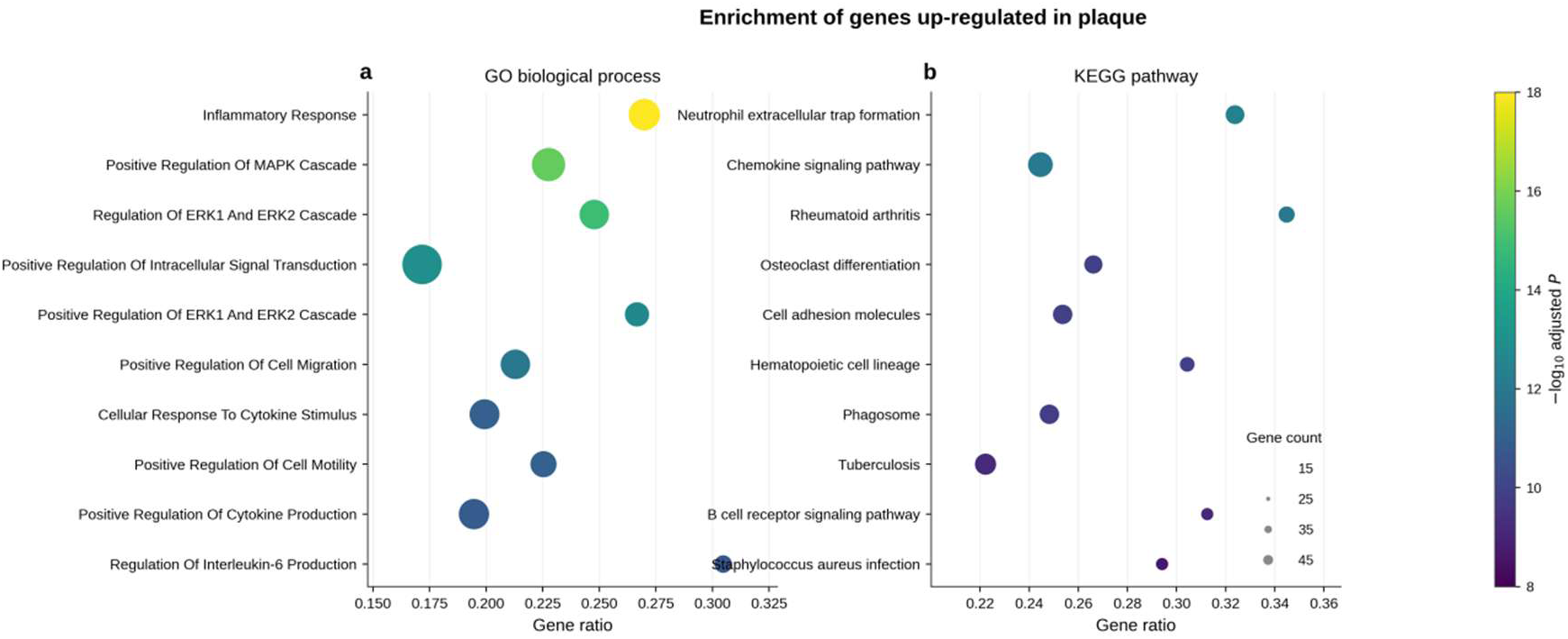
Functional enrichment of genes upregulated in plaque. Top 10 Gene Ontology biological processes (left) and KEGG pathways (right) ranked by Benjamini– Hochberg-adjusted P. Enrichment is dominated by inflammatory and myeloid programmes rather than copper- or metabolism-specific terms.

To connect the copper genes to this inflammatory signature at the sample level, we correlated per-sample gene expression with ssGSEA immune-cell infiltration scores (64 samples; 32 plaque, 32 intact). The copper importers tracked strongly and positively with myeloid/inflammatory infiltration: *SLC31A2* correlated with dendritic-cell (Pearson r = +0.91), M1-macrophage (+0.87), M2-macrophage (+0.86) and neutrophil (+0.83) scores, and *SLC31A1* showed the same pattern (M1 r = +0.78). Strikingly, the copper *exporter ATP7B* correlated *negatively* with the same infiltrating populations (M1-macrophage r = −0.73), as did *GLS* (r = −0.64) and *PDHA1* (r = −0.65) (**Figure 3**).

**Figure 3.**
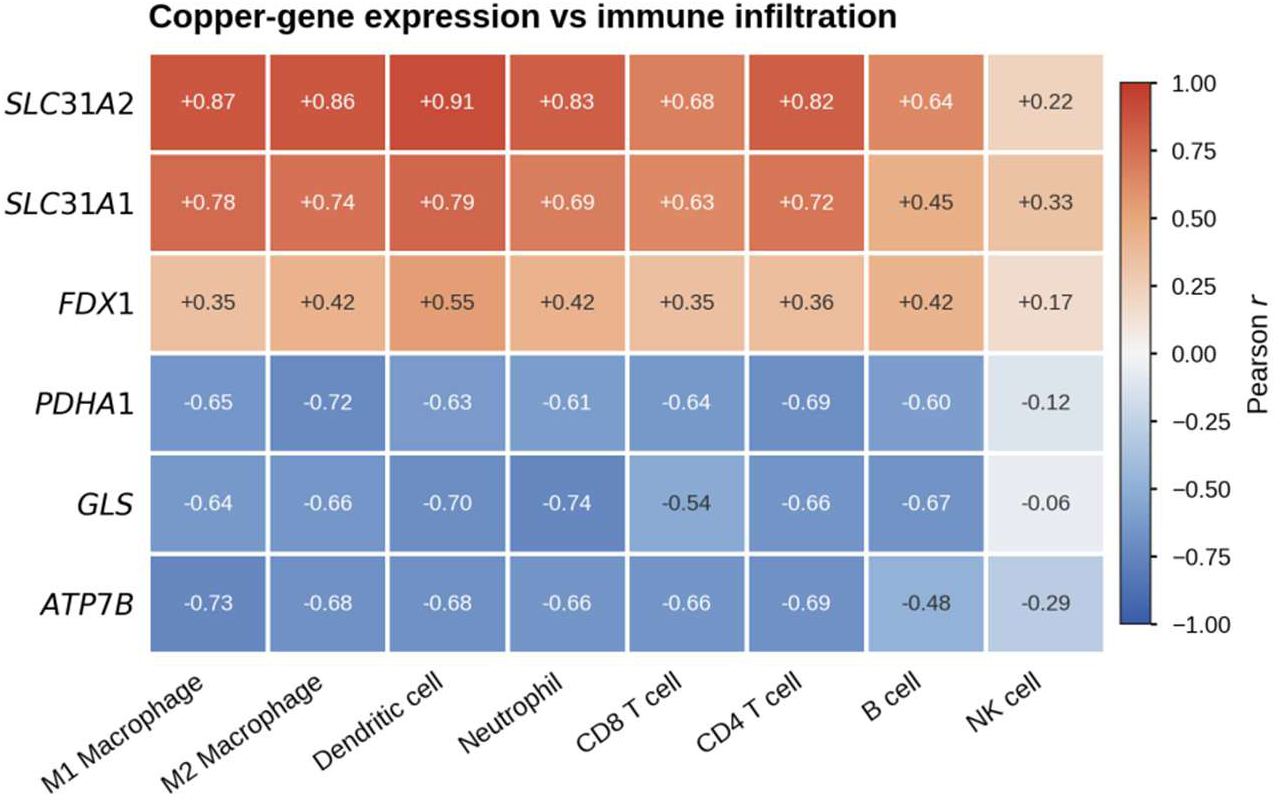
Copper-gene expression tracks immune-cell infiltration. Pearson correlation between per-sample expression of copper/cuproptosis genes and ssGSEA immune-cell infiltration scores (GSE43292, n = 64 samples). Copper importers (*SLC31A2*, *SLC31A1*) correlate positively with myeloid populations, whereas the exporter *ATP7B*, *GLS* and *PDHA1* correlate negatively.

Thus, in our hands and across two cohorts, the plaque-associated dysregulation of copper-handling genes is real and reproducible—but it is tightly coupled to the degree of inflammatory and myeloid-cell infiltration of the lesion, with copper *importers* rising and the *exporter* falling as infiltration increases. This pattern is that expected of markers of the inflamed plaque microenvironment, and it motivates—but does not by itself establish—a causal test, which we perform next.

### 3.2 The MR pipeline is validated by the positive control *LPL*

Using four independent whole-blood instruments, higher genetically predicted *LPL* expression was strongly associated with lower CAD risk (IVW OR 0.90 per SD, 95% CI 0.87–0.93, P = 4.1 × 10^−12^), with concordant estimates across MR-Egger, weighted median, and weighted mode, and no evidence of heterogeneity (Q P = 0.60) or directional pleiotropy (Egger intercept P = 0.45). This recovers the established causal, protective relationship of *LPL* with coronary disease and confirms that the eQTLGen→CAD pipeline (Z-to-β conversion, clumping, harmonization, MR) behaves correctly. The positive control was intended to validate the direction and harmonization of the MR pipeline, not to demonstrate that single-causal-variant colocalization must succeed at a known multi-signal locus; indeed, as noted below, *LPL* does not itself colocalize under the single-variant model.

### 3.3 No cuproptosis/copper gene shows colocalization-supported causal evidence for CAD

Across the 11 cuproptosis/copper genes with usable instruments, MR estimates were null for the well-instrumented genes and none was supported by colocalization (all PP.H4 < 0.02; **Table 2**, **Table 3**). Two genes reached nominal MR significance, but only in single-instrument (Wald-ratio) analyses:

- ***MTF1*** (OR 1.16 per SD, 95% CI 1.05–1.28, P = 2.7 × 10⁻^3^) — colocalization strongly refuted a shared variant (**PP.H3 = 0.99998, PP.H4 = 3.6 × 10⁻^6^**). Inspection of the region makes the reason concrete (**Figure 6**): the region does contain a genuine, genome-wide-significant CAD signal (lead variant rs61776719, −log₁₀P = 11.2), but it lies ∼120 kb away from the lead *MTF1* eQTL (rs3748804, −log₁₀P = 30.3). The two variants are correlated through linkage disequilibrium but distinct, so a single-instrument analysis misattributes the neighbouring disease signal to *MTF1* expression.
- ***DLAT*** (OR 1.41 per SD, 95% CI 1.13–1.75, P = 2.2 × 10⁻^3^) — not colocalized (**PP.H4 = 0.29**); here both signals are weak (lead eQTL −log₁₀P = 7.5; lead CAD variant −log₁₀P = 4.5, not genome-wide significant) and the lead variants are ∼180 kb apart, so the data are uninformative rather than positively against a shared variant.

**Table 2.** Mendelian randomization estimates for cuproptosis/copper genes and the positive control on CAD (eQTLGen whole-blood instruments).

| Gene | Class | N instruments | Primary method | OR per SD (95% CI) | P |
| --- | --- | --- | --- | --- | --- |
| ATP7B | copper transport | 3 | IVW | 1.009 (0.972–1.048) | 0.62 |
| SLC31A2 | copper transport | 3 | IVW | 1.020 (0.982–1.059) | 0.31 |
| SLC31A1 | copper transport | 3 | IVW | 0.996 (0.955–1.040) | 0.86 |
| FDX1 | cuproptosis | 2 | IVW | 0.968 (0.937–1.001) | 0.054 |
| LIPT1 | cuproptosis | 3 | IVW | 0.987 (0.955–1.020) | 0.43 |
| LIAS | cuproptosis | 3 | IVW | 1.017 (0.979–1.057) | 0.39 |
| GLS | cuproptosis | 2 | IVW | 0.985 (0.945–1.026) | 0.46 |
| DLD | cuproptosis | 1 | Wald ratio | 1.042 (0.932–1.164) | 0.47 |
| DLST | cuproptosis | 1 | Wald ratio | 0.989 (0.921–1.062) | 0.76 |
| DBT | cuproptosis | 1 | Wald ratio | 0.921 (0.741–1.144) | 0.46 |
| CDKN2A | cuproptosis | 2 | IVW | 0.528 (0.041–6.87) | 0.63 |
| MTF1 | cuproptosis | 1 | Wald ratio | **1.161 (1.053–1.280)** | **2.7 × 10<sup>-3</sup>** |
| DLAT | cuproptosis | 1 | Wald ratio | **1.409 (1.132–1.754)** | **2.2 × 10<sup>-3</sup>** |
| **LPL (positive control)** | lipid | 4 | IVW | **0.897 (0.870–0.925)** | **4.1 × 10<sup>-12</sup>** |

Bonferroni threshold $P = 3.6 \times 10^{-3}$ . MTF1 and DLAT are single-instrument nominal associations refuted by colocalization (Table 3).

**Table 3.** Colocalization of gene eQTL and CAD signals.

| Gene | SNPs | PP.H3 (distinct variants) | PP.H4 (shared variant) | Interpretation |
| --- | --- | --- | --- | --- |
| DLAT | 6,048 | 0.27 | 0.29 | not colocalized |
| DBT | 5,970 | 0.11 | 0.010 | not colocalized |
| LIAS | 7,281 | 0.17 | 0.004 | not colocalized |
| GLS | 5,473 | 0.24 | 0.003 | not colocalized |
| LIPT1 | 5,464 | 0.12 | 0.002 | not colocalized |
| SLC31A2 | 6,887 | 0.52 | 0.001 | not colocalized |
| SLC31A1 | 6,324 | 0.46 | 0.001 | not colocalized |
| ATP7B | 5,458 | 0.69 | 0.001 | not colocalized |
| LPL (control) | 9,429 | 0.9998 | $2.2 \times 10^{-4}$ | see note |
| FDX1 | 6,067 | 0.986 | $2.0 \times 10^{-4}$ | not colocalized |
| MTF1 | 6,358 | 0.99999 | $3.6 \times 10^{-6}$ | not colocalized |
| DLD | 5,865 | 0.9999 | $4.0 \times 10^{-7}$ | not colocalized |
| DLST | 6,080 | 0.99999 | $1.2 \times 10^{-7}$ | not colocalized |
| CDKN2A | 7,455 | 1.00 | $8.5 \times 10^{-16}$ | not colocalized |

*Note: LPL shows a strong, robust MR effect yet PP.H3 dominates. This reflects allelic heterogeneity in the LPL locus, which violates coloc's single-causal-variant assumption; it illustrates that colocalization can fail to confirm a genuinely causal gene in multi-signal regions, and argues for interpreting MR and colocalization jointly (see Discussion).*

Because single-instrument cis-MR cannot separate the causal effect of the target gene from that of a neighbouring gene in linkage disequilibrium, and because colocalization did not support a shared causal variant for either gene, these two associations are most parsimoniously explained by LD confounding rather than a causal effect of *MTF1* or *DLAT* expression.

For the copper transporters and *FDX1*—the genes of primary biological interest— estimates were null across all MR methods and not colocalized: *SLC31A2* IVW OR 1.02 (0.98–1.06), *ATP7B* 1.01 (0.97–1.05), *SLC31A1* 1.00 (0.95–1.04), *FDX1* 0.97 (0.94–1.00, P = 0.054). Sensitivity analyses showed no directional pleiotropy for any gene (all Egger-intercept P > 0.28); modest heterogeneity was detected only for *SLC31A1* (Q P = 0.045), for which the robust weighted-median estimate remained null.

Regional and colocalization results are shown in **Figures 4–6**.

**Figure 4.**
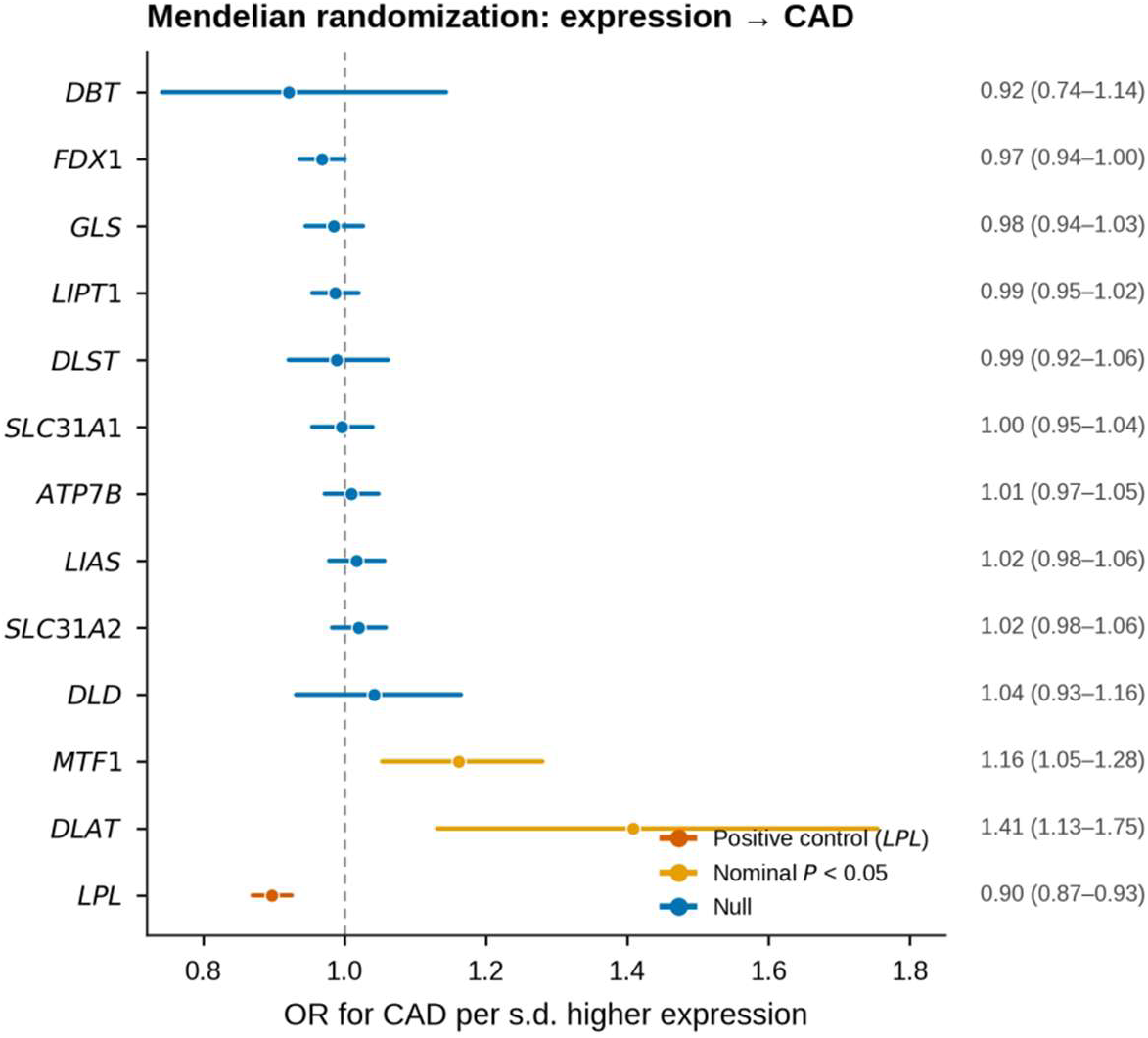
Mendelian randomization estimates for gene expression on coronary artery disease. Odds ratio (95% CI) for CAD per standard-deviation higher genetically predicted expression, using eQTLGen whole-blood instruments. Red, positive control *LPL*; orange, nominal P < 0.05; blue, null. Numbers of instruments are given at right. *CDKN2A* is omitted for scale (OR 0.53, 95% CI 0.04–6.87; uninformative instruments) and is reported in Table 2.

**Figure 5.**
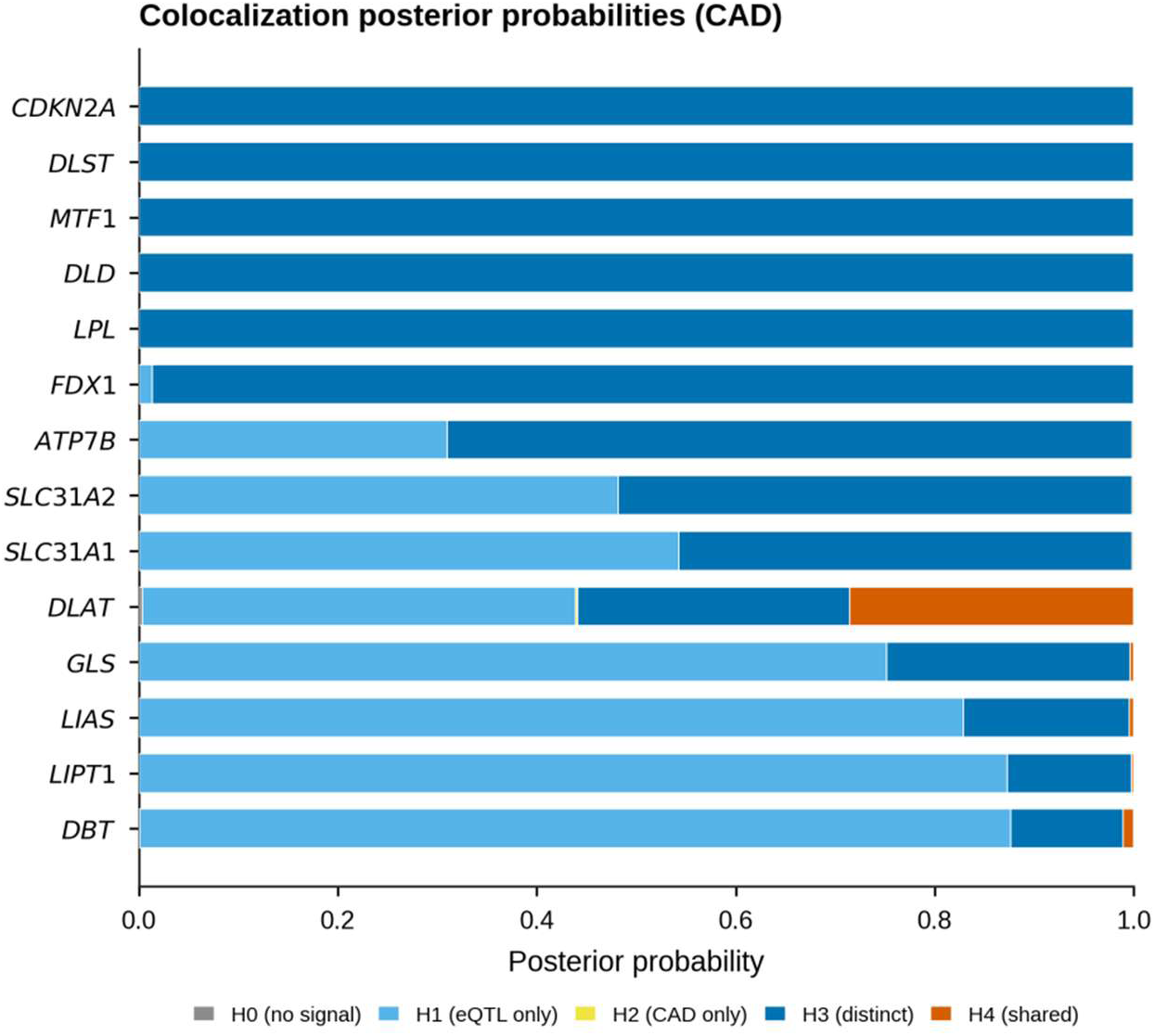
Colocalization posterior probabilities. Stacked posterior probabilities for the five coloc hypotheses for each gene (eQTL vs. CAD). No gene reaches PP.H4 > 0.7. The two nominally significant MR genes, *MTF1* and *DLAT*, are dominated by PP.H3 (distinct causal variants), indicating linkage-disequilibrium confounding rather than causality.

**Figure 6.**
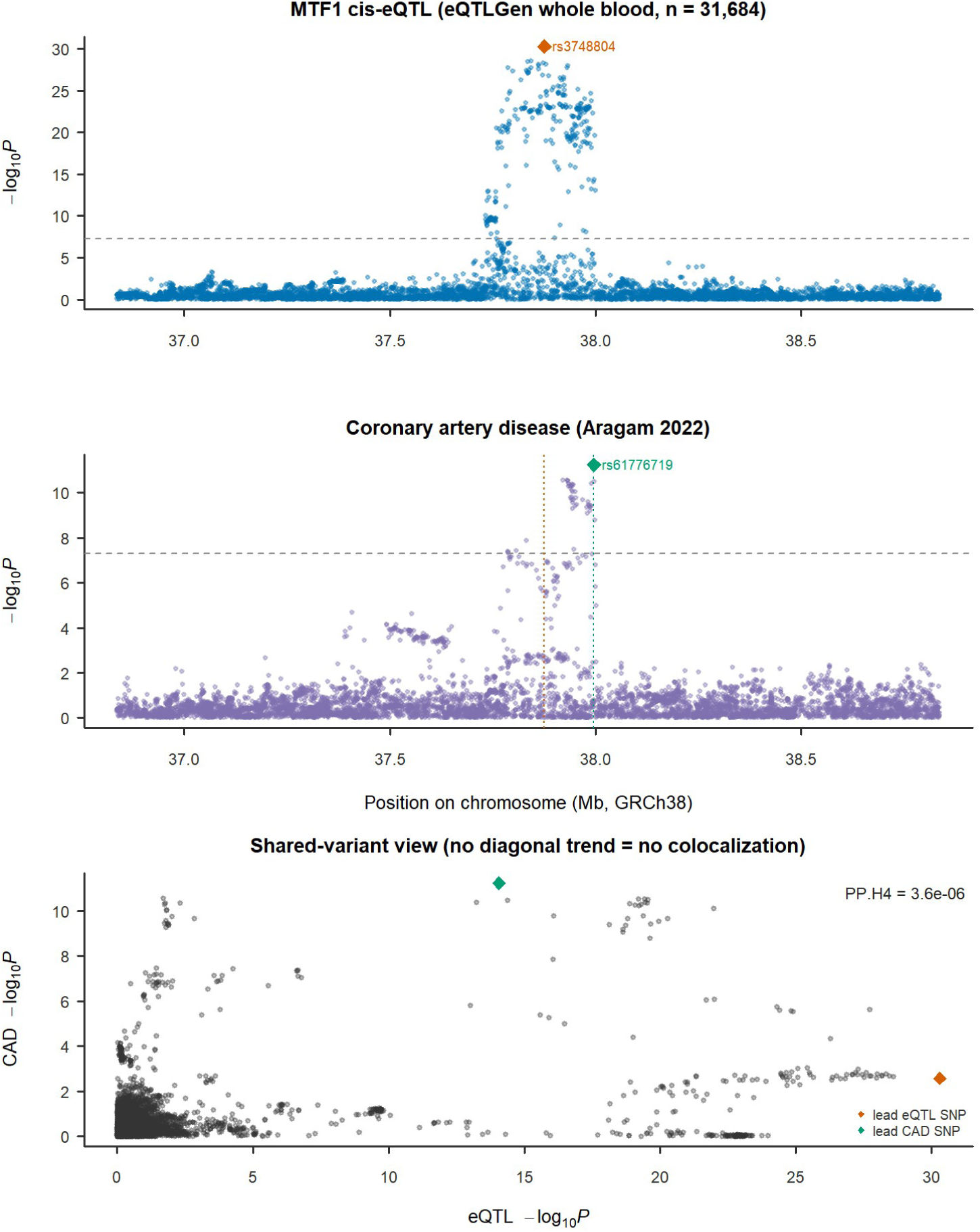
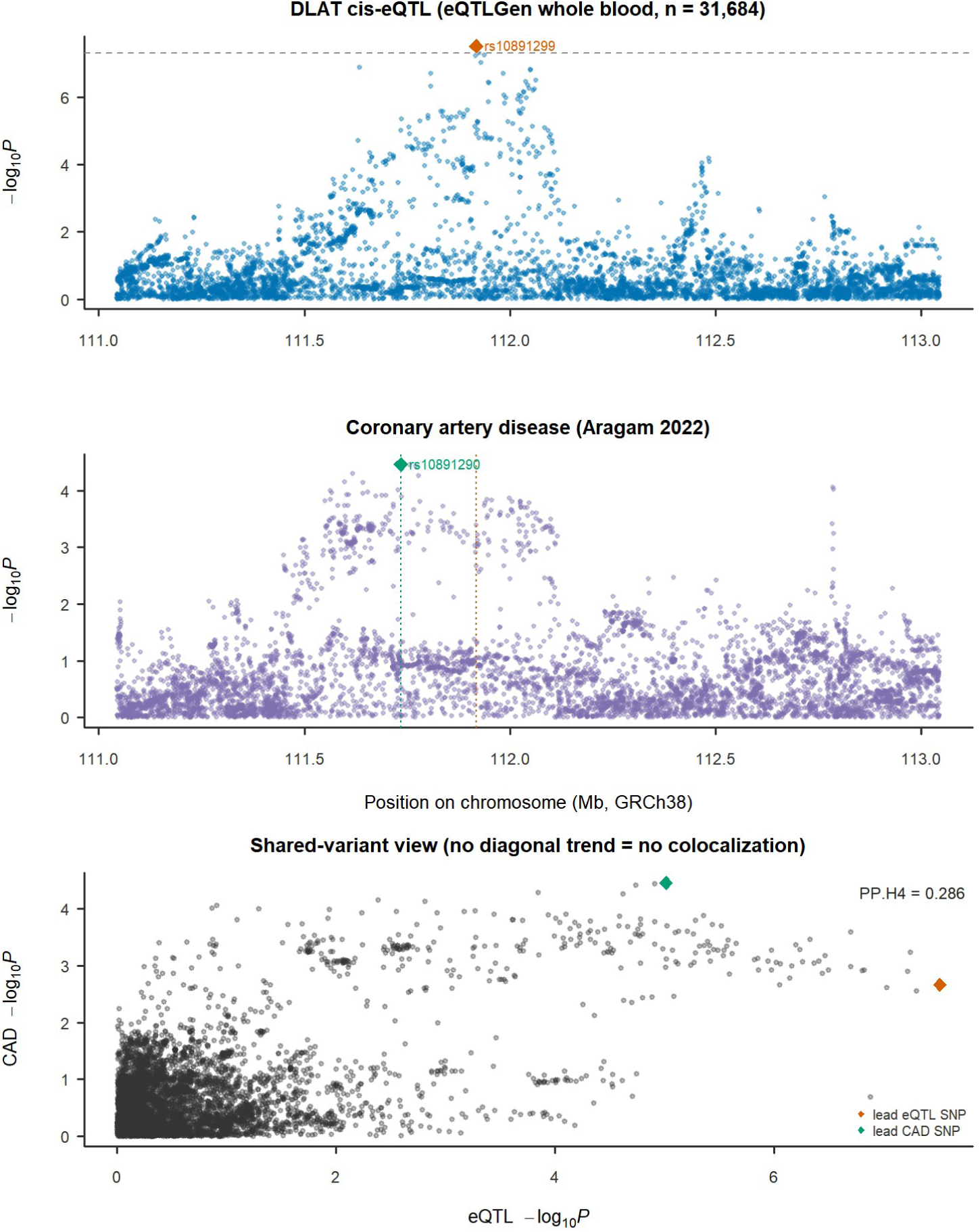
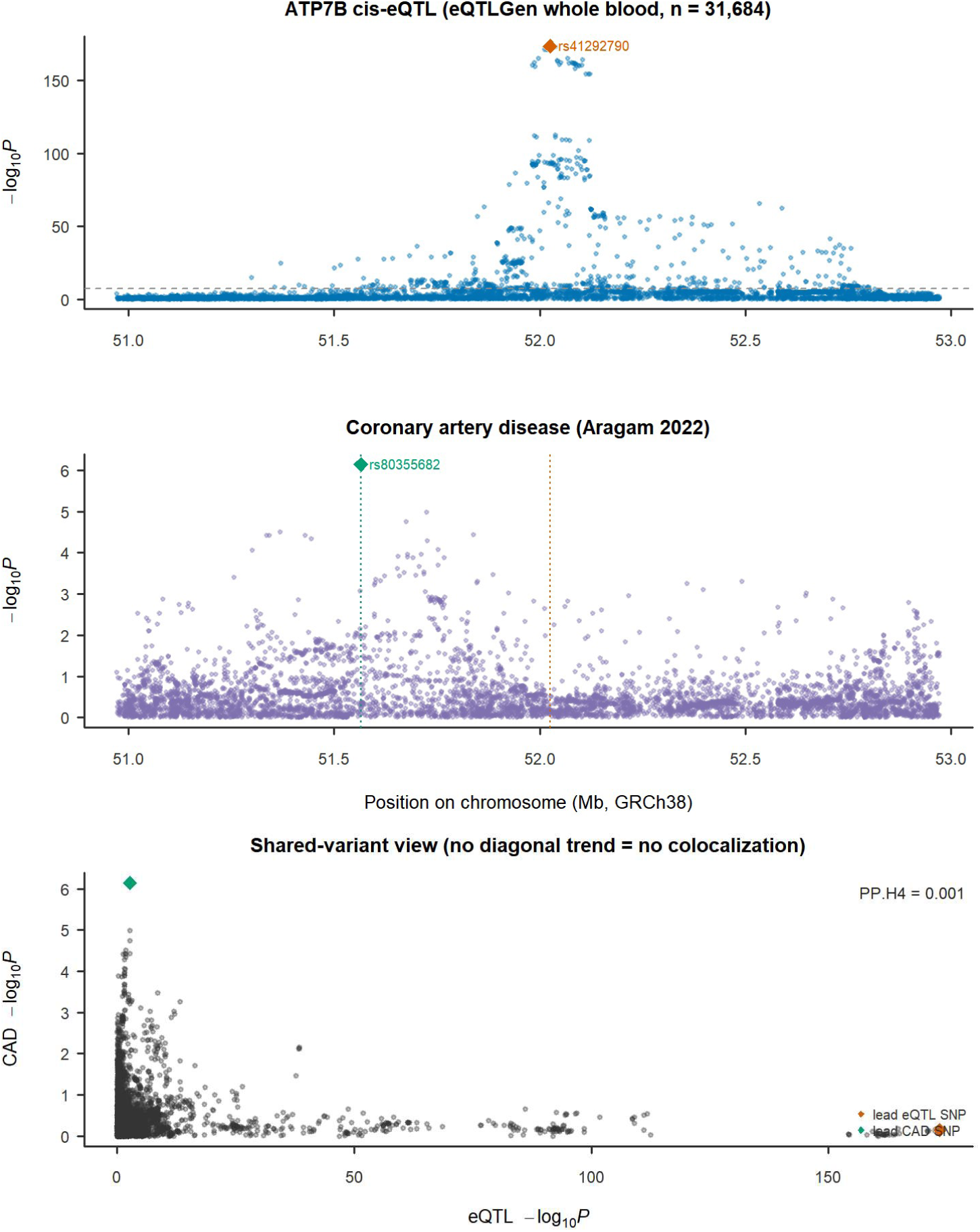
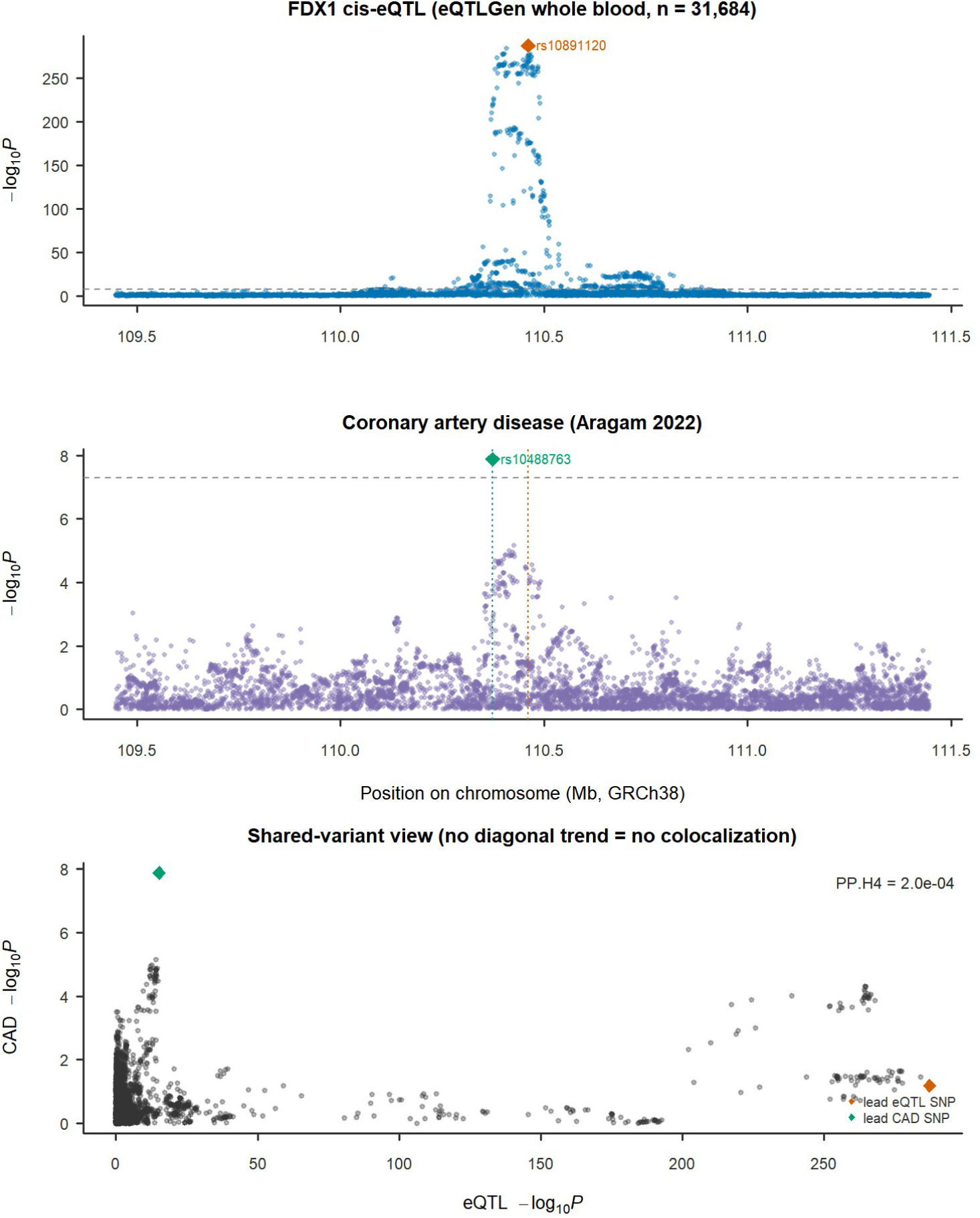
Regional comparison of eQTL and CAD association signals. For *MTF1*, *DLAT*, *ATP7B* and *FDX1*: cis-eQTL −log₁₀P (top) and CAD −log₁₀P (middle) plotted against GRCh38 position (variants matched by rsID), and the corresponding shared-variant scatter (bottom). Lead eQTL and lead CAD variants are marked by diamonds and dotted vertical lines. In every locus the two lead variants are separated (e.g. ∼120 kb for *MTF1*, ∼180 kb for *DLAT*) and the scatter shows no diagonal trend, illustrating the absence of colocalization. The *MTF1* panel is the clearest example: a genuine genome-wide-significant CAD signal (rs61776719, −log₁₀P = 11.2) sits ∼120 kb from the lead *MTF1* eQTL (rs3748804, −log₁₀P = 30.3), explaining why single-instrument Mendelian randomization produced a spurious association.

### 3.4 The analysis is adequately powered for the well-instrumented genes

For genes with multiple strong instruments, the study had 80% power at α = 0.05 to detect modest effects: the minimum detectable OR per SD of expression was ≈1.045–1.063 for *LPL*, *FDX1*, *LIPT1*, *ATP7B*, *SLC31A2*, *SLC31A1*, *LIAS* and *GLS* (**Table 4**). Thus, for the copper transporters and *FDX1*, the null results exclude anything but very small causal effects and are not attributable to insufficient power. Single-instrument genes (*DBT*, *DLAT*, *DLST*, *MTF1*, *DLD*) were less well powered (minimum detectable OR 1.11–1.37), and *CDKN2A* was effectively uninformative (minimum detectable OR ≈39; weak, imprecise instruments).

**Table 4.** Statistical power (minimum detectable OR per SD of expression at 80% power, α = 0.05).

| Gene | N instruments | Min. detectable OR/SD |
| --- | --- | --- |
| LPL | 4 | 1.045 |
| FDX1 | 2 | 1.048 |
| LIPT1 | 3 | 1.048 |
| ATP7B | 3 | 1.055 |
| SLC31A2 | 3 | 1.055 |
| LIAS | 3 | 1.056 |
| GLS | 2 | 1.061 |
| SLC31A1 | 3 | 1.063 |
| DLST | 1 | 1.107 |
| MTF1 | 1 | 1.150 |
| DLD | 1 | 1.173 |
| DBT | 1 | 1.363 |
| DLAT | 1 | 1.368 |
| CDKN2A | 2 | 39.1 (uninformative) |

### 3.5 Secondary outcome and secondary tissue

Using the same whole-blood instruments, Mendelian randomization for large-artery atherosclerotic stroke was null for every gene tested: *ATP7B* OR 1.13 (95% CI 0.91–1.40, P = 0.26), *SLC31A2* OR 0.99 (0.82–1.20, P = 0.94), *SLC31A1* OR 0.92 (0.83–1.02, P = 0.12), *FDX1* OR 0.98 (0.76–1.27, P = 0.88), *LIPT1* OR 0.94 (0.83–1.07, P = 0.37), *GLS* OR 1.14 (0.95–1.36, P = 0.15) (**Table S2**). Colocalization likewise showed no shared causal variant (all PP.H4 ≤ 0.053; **Table S3**). These analyses should not, however, be read as independent confirmation: with only 6,399 cases the stroke GWAS carried essentially no association signal in any of the cis regions examined (regional minimum P ≈ 10⁻^3^), and the confidence intervals are correspondingly wide—for example, the *ATP7B* interval cannot exclude a 40% increase in risk. The secondary outcome therefore adds breadth rather than evidence, and our inference rests on the well-powered CAD analysis.

The parallel tibial-artery (GTEx v8, n = 584) analysis was limited by the small eQTL sample to a single independent instrument per gene, permitting only Wald-ratio estimates; these were likewise null (e.g. *ATP7B* OR 1.003, 95% CI 0.988–1.017, P = 0.74; *SLC31A2* OR 0.98, 0.94–1.02, P = 0.27), and colocalization against CAD again showed no shared causal variant for any gene (all PP.H4 ≤ 0.015; **Table S4**). Notably, in this arm the *FDX1* region produced a decisive PP.H3 of 0.986, confirming that the genome-wide-significant CAD signal in that region (minimum P = 1.3 × 10⁻^8^) is driven by a variant distinct from the *FDX1* eQTL.

## 4. Discussion

In a two-sample cis-Mendelian randomization and colocalization study anchored on a well-powered European CAD GWAS, we found no colocalization-supported evidence that the genetically determined expression of any cuproptosis- or copper-transport gene is causally associated with coronary artery disease. The positive control *LPL* reproduced its known protective effect (OR 0.90, P = 4 × 10⁻^12^), confirming that the analytic pipeline can detect a true causal signal of modest size. For the copper transporters and the cuproptosis regulator *FDX1*, the null estimates were precise enough to exclude odds ratios larger than about 1.05 per SD of expression, so they cannot be attributed to insufficient power.

Two genes, *MTF1* and *DLAT*, met the nominal Bonferroni MR threshold in single-instrument (Wald-ratio) analyses but were not credible after colocalization.^28^ Colocalization, however, refuted a shared causal variant in both cases—most emphatically for *MTF1* (PP.H3 = 0.99998)—and regional inspection supplies the physical explanation: in the *MTF1* locus a real, genome-wide-significant CAD association sits roughly 120 kb from the lead *MTF1* eQTL, close enough to be correlated through linkage disequilibrium but plainly a different variant (Figure 6). This is the classic failure mode of single-instrument cis-MR, in which the instrument tags a nearby causal variant belonging to a different gene, and it illustrates why colocalization is an essential adjunct to cis-MR rather than an optional add-on. Had we reported the MR alone, *MTF1* would have been presented as a novel cuproptosis-related CAD gene. Interpreted together, MR and colocalization concur that neither association is a credible causal effect.

Our positive control also carries a methodological lesson in the opposite direction: *LPL* was strongly causal by MR yet failed colocalization (PP.H3 dominant), because the *LPL* locus harbours multiple independent regulatory signals that violate the single-causal-variant assumption of the coloc.abf model. This means a null colocalization result alone should not be over-interpreted as excluding causality; it is the *concordance* of null MR and null colocalization across genes—together with adequate power—that underpins our conclusion.

These genetic findings are consistent with the broader evidence on copper and cardiovascular disease. Prior MR studies of circulating copper have reported null or inverse (protective) associations with coronary disease (copper→CAD OR 0.92 and copper→ischaemic heart disease OR 0.94),^29,30^ rather than the harmful effect that would be predicted if plaque copper accumulation were causally atherogenic. Our results extend this from the level of the copper *phenotype* to the level of individual copper-handling and cuproptosis *genes*, and reach the same conclusion. They also speak directly to the descriptive literature: the genes most often nominated from plaque transcriptomes—*SLC31A1*, *SLC31A2*, *FDX1*, *GLS*, *CDKN2A*^3,4,5^—are precisely those for which we find no causal or colocalization support, including in the carotid tissue from which several of those signatures were derived.^5^

How, then, should the well-replicated upregulation of these genes in plaque—which we also observed across two cohorts—be understood? Our own transcriptomic data point to the answer. The plaque-upregulated gene set was enriched overwhelmingly for inflammatory and myeloid programs (inflammatory response, neutrophil extracellular trap formation, chemokine signaling), and at the single-sample level the copper importers *SLC31A2* and *SLC31A1* rose in tight proportion to macrophage, dendritic-cell and neutrophil infiltration (*SLC31A2* vs. M1-macrophage r = +0.87), while the copper exporter *ATP7B* fell (r = −0.73). This is precisely the signature expected if copper-import gene expression is a **marker of inflammatory-cell infiltration of the lesion**, tracking the myeloid compartment that expands during plaque progression,^31^ rather than a **cause** of that progression. The most parsimonious synthesis of the observational and causal data is therefore that the plaque upregulation of these genes is a *consequence* of the inflamed, macrophage-rich plaque microenvironment—not a driver of disease initiation or progression. This reconciles the strong observational signal with the null causal signal, and carries a direct practical implication: candidate targets nominated purely from differential-expression analyses of diseased tissue, however reproducible, may be bystanders of infiltration and require causal validation before therapeutic pursuit—in contrast to inflammation itself, for which causal human genetic and randomized-trial evidence exists.^32^

## Limitations

First, and most importantly, cis-eQTL MR tests only the effect of *germline-determined, baseline* variation in gene expression. It cannot capture the potentially disease-relevant biology of copper accumulation and cuproptosis *within* the plaque microenvironment, which is a local, acquired, non-germline process that no common eQTL indexes. Our null therefore does **not** disprove a role for cuproptosis in plaque biology; it shows specifically that common genetic variation in the expression of these genes does not measurably alter lifelong CAD risk. Mendelian randomization estimates a lifelong, germline-determined expression effect, whereas cuproptosis may be triggered locally by copper accumulation, shear stress, oxidative stress or mitochondrial state—acquired processes that common cis-eQTLs need not capture.^33,34^ Second, the whole-blood eQTL analysis tests systemically, genetically regulated expression, whereas plaque upregulation may reflect cell-composition shifts and local disease-state induction that a blood cis-eQTL does not capture; although blood is the tissue in which several of these genes (including *SLC31A2*) are most highly expressed, tissue-specific regulation in the arterial wall, endothelium, macrophages or smooth muscle may differ, and the small GTEx arterial sample limited the parallel tissue analysis to one instrument per gene. Third, some genes (*PDHA1*, *ATP7A*) are X-linked or not expressed in blood and could not be instrumented in eQTLGen; *PDHA1* additionally could not be tested against the autosome-only CAD GWAS. Fourth, colocalization used the single-causal-variant model, with the caveat discussed above. Fifth, the analysis was restricted to European-ancestry data to match instrument and outcome populations, limiting generalizability.

**Strengths** include the use of two complementary causal methods (MR and colocalization) with a validated positive control, a large and well-powered outcome dataset, explicit power/precision analysis, and full reproducibility from public summary statistics.

## 5. Conclusion

In available eQTL tissues, the expression of cuproptosis- and copper-transport genes as determined by common cis-regulatory variants did not support a detectable causal effect on coronary artery disease risk. This does not exclude a role for locally acquired cuproptosis within the plaque; their upregulation in atherosclerotic tissue is best interpreted as a marker or consequence of disease rather than a germline-encoded driver. These results argue for causal validation before cuproptosis-pathway genes are advanced as therapeutic targets in atherosclerosis on the basis of expression studies alone.

## Data and code availability

All summary statistics are public: eQTLGen (https://www.eqtlgen.org), GTEx v8, CAD GCST90132314, LAS GCST90104542; carotid transcriptomes GSE43292 and GSE28829. Analysis scripts (data download, instrument construction, MR, colocalization, power) are available at Zenodo, DOI: 10.5281/zenodo.21615076, and at GitHub (github.com/wguan964-jpg/cuproptosis-mr-cad).

## Research Perspective

What New Question Does This Study Raise?

If common cis-regulatory variation in cuproptosis and copper-transport genes does not support a detectable causal effect on coronary artery disease risk, what plaque-local or disease-state-specific mechanisms explain their consistent upregulation in atherosclerotic lesions, and do these signals primarily reflect inflammatory-cell infiltration or cell-state changes rather than germline-regulated expression?

What Question Should Be Addressed Next?

Cell-type-specific studies using single-cell multiomics, spatial transcriptomics, and experimental perturbation models are needed to determine whether copper accumulation and cuproptosis-like mitochondrial injury occur within defined plaque cell populations, such as macrophages, foam cells, or stressed vascular cells, and whether they contribute causally to plaque progression or instability before cuproptosis-pathway genes are prioritized as therapeutic targets.

## Author Contributions

G.W. and W.L. contributed equally to this work. G.W., W.L., and Y.X. designed the study. G.W. and W.L. performed the statistical analyses. G.W., W.L., and M.G. analyzed the transcriptomic data. G.W. drafted the manuscript. Y.X. supervised the project. All authors critically revised the manuscript and approved the final version.

## Ethical Statement

This study used only publicly available summary-level genetic data and does not involve individual-level human subjects research. No institutional review board approval was required.

## Trial Registration

Not applicable (this is not a clinical trial).

---

**Table S1 (supplement). Heterogeneity and pleiotropy tests** (from eqtlgen_sensitivity.csv): Cochran’s Q P-values — ATP7B 0.64, SLC31A2 0.62, SLC31A1 0.045, LIPT1 0.27, LIAS 0.11, LPL 0.60; MR-Egger intercept P-values all > 0.28 (no directional pleiotropy).

**Table S2 (supplement).** **Mendelian randomization for large-artery atherosclerotic stroke** (eQTLGen whole-blood instruments; GCST90104542, 6,399 cases / 1,234,808 controls).

| Gene | N instruments | Method | OR per SD (95% CI) | P |
| --- | --- | --- | --- | --- |
| ATP7B | 3 | IVW | 1.130 (0.913–1.399) | 0.26 |
| SLC31A2 | 3 | IVW | 0.993 (0.820–1.202) | 0.94 |
| SLC31A1 | 3 | IVW | 0.919 (0.827–1.022) | 0.12 |
| FDX1 | 2 | IVW | 0.981 (0.757–1.272) | 0.88 |
| LIPT1 | 3 | IVW | 0.942 (0.827–1.073) | 0.37 |
| GLS | 2 | IVW | 1.139 (0.953–1.360) | 0.15 |
| DBT | — | — | instrument absent from stroke GWAS | — |

**Table S3 (supplement).** **Colocalization of whole-blood eQTL and large-artery atherosclerotic stroke signals** (eQTLGen, n = 31,684; all cis-region variants; GCST90104542).

| Gene | SNPs | PP.H1 | PP.H3 | PP.H4 |
| --- | --- | --- | --- | --- |
| DBT | 3,502 | 0.823 | 0.116 | 0.053 |
| GLS | 3,266 | 0.848 | 0.122 | 0.031 |
| ATP7B | 3,260 | 0.892 | 0.092 | 0.016 |
| SLC31A1 | 4,597 | 0.836 | 0.150 | 0.014 |
| LIPT1 | 3,133 | 0.908 | 0.078 | 0.014 |
| FDX1 | 3,982 | 0.847 | 0.142 | 0.012 |
| SLC31A2 | 4,580 | 0.850 | 0.140 | 0.010 |

*No gene reaches PP.H4 > 0.7. The dominance of PP.H1 (eQTL signal present, outcome signal absent) reflects the limited power of the stroke GWAS in these regions rather than positive evidence against a shared variant. Genes are restricted to those with extracted stroke cis-regions.*

**Table S4 (supplement).** **Colocalization of tibial-artery eQTL and CAD signals** (GTEx v8, n = 584; all cis variants, palindromic SNPs retained).

| Gene | SNPs | PP.H1 | PP.H3 | PP.H4 |
| --- | --- | --- | --- | --- |
| FDX1 | 6,002 | 0.014 | **0.986** | $3.8 \times 10^{-5}$ |
| ATP7B | 4,990 | 0.313 | 0.685 | 0.0013 |
| SLC31A2 | 6,550 | 0.488 | 0.508 | 0.0033 |
| SLC31A1 | 6,553 | 0.533 | 0.459 | 0.0018 |
| GLS | 5,437 | 0.746 | 0.239 | 0.0150 |
| DBT | 6,134 | 0.865 | 0.129 | 0.0064 |
| LIPT1 | 5,101 | 0.883 | 0.114 | 0.0023 |

